# Predictive modeling for bacterial vaginosis in a Tanzanian cohort of women living with HIV

**DOI:** 10.64898/2026.03.11.26347251

**Authors:** Diandra P. Ojo, Wambui Gachunga, Carleigh C. Sokolik, Ivana K. Parker

## Abstract

**Introduction:** The vaginal microbiome is an important factor affecting both HIV risk and reproductive health in women living with HIV, and women of African descent are disproportionately affected. Here we use 16S rRNA sequencing data to predict BV in HIV-positive women living in Africa and delineate microbial features that are important for accurate prediction in this cohort compared to two HIV negative cohorts.

**Methods:** Our study population was comprised of a cohort of HIV-positive women living in Tanzania, and two cohorts of HIV-negative women living in the United States. Using OTU variables from data in the Tanzanian cohort, we used random forest, logistic regression, support vector machine, and multi-layer perceptron algorithms to predict BV outcome. To evaluate our models, we compared predicted BV outcome to BV outcome determined using Nugent scoring.

**Results:** In evaluating model performance, all four models predicted BV outcome better for the HIV-negative cohorts than the HIV-positive cohorts. Overall, models were better at predicting negative BV outcomes for low Nugent scores than positive BV outcomes for high Nugent scores. Evaluation of significant predictor variables of BV in each cohort revealed that shared features existed between the Tanzanian and Symptomatic cohorts, but not among all three cohorts. Upon comparing performance of models in predicting BV outcome for Black women in HIV-positive and HIV-negative cohorts, we observed that all four models perform better at predicting BV in the HIV-negative cohorts.

**Conclusions:** The lower predictive performance observed for this Tanzanian HIV positive cohort, coupled with the difference in microbial communities important for accurate prediction suggest that distinct microbial communities in women who are living with HIV may create challenges that affect the accuracy of BV diagnosis and treatment. This study highlights the need for diagnostic tools that consider unique biological and epidemiological factors of populations to address health disparities.

## Introduction

HIV remains a relentless global health crisis, disproportionately affecting women of African descent. In sub-Saharan Africa, adolescent girls and young women accounted for over 77% of new infections among 15 to 24 year olds^1^. This region bears the brunt of the epidemic, with women representing nearly 60% of all new HIV infections ^2^. The interplay between vaginal health and HIV transmission is increasingly recognized, with bacterial vaginosis (BV) emerging as a significant factor^3^. BV is a prevalent vaginal condition among women of reproductive age, characterized by an imbalance in the vaginal microbiota where Lactobacillus-dominated flora is replaced by a polymicrobial community of anaerobes^4^. BV presents symptoms such as vaginal discharge, a fishy odor, vaginal itching, and burning during urination, though many women may remain asymptomatic. BV is also associated with several complications, including an increased risk of sexually transmitted infections (STIs), pelvic inflammatory disease (PID), and adverse pregnancy outcomes such as preterm birth and low birth weight^3,5^. Recent data indicate that BV prevalence ranges from 23% to 29%, with sexually active women being predominantly affected^6^. In Tanzania, a study conducted in Dar es Salaam found a significant prevalence in that 33.2% of women with vaginal discharge had BV^7^.

Elevated inflammation in the female genital tract is linked to increased HIV risk, with diverse anaerobic bacterial communities significantly heightening this risk by inducing mucosal HIV target cells. Conversely, *Lactobacillus spp* are typically associated with reduced inflammation and a lower risk of HIV acquisition ^8^. Diagnosis of BV relies on Amsel’s criteria^9^ or Nugent score, a Gram stain-based scoring system^10^. However, the variability in vaginal microbiota profiles across different ethnic groups can impact the diagnostic accuracy of BV. Women of African ancestry often have distinct vaginal microbial communities with a higher prevalence of BV-associated bacteria and lower levels of Lactobacillus species^11,12^. These microbial profiles are often classified into community state types (CSTs), which reflect the dominant bacterial species within the vaginal microbiome: CST I is dominated by *Lactobacillus crispatus*, CST II by *Lactobacillus gasseri*, CST III by *Lactobacillus iners*, CST V by *Lactobacillus jensenii*, and CST IV is characterized by a lack of *Lactobacillus* dominance and instead comprises a diverse mix of anaerobic bacteria^12^. CST distribution has been shown to vary by race, with Black women more frequently exhibiting a non-*Lactobacillus*-dominated CST which is associated with increased inflammation and a heightened risk of bacterial vaginosis^13^. Furthermore, socioeconomic and cultural barriers further complicate the diagnosis and treatment of BV in women of African ancestry. These women often face significant barriers to healthcare access, including socioeconomic factors, geographic limitations, and systemic healthcare inequalities, which can delay diagnosis and treatment^14,15^.

Advances in sequencing technology and artificial intelligence can improve our understanding and management of bacterial vaginosis (BV) by revealing intricate microbial shifts in the vaginal environment. Leveraging sequencing data from vaginal fluid, machine learning tools have become pivotal in identifying patterns and predicting outcomes associated with BV. Recent studies have utilized machine learning algorithms to analyze 16S sequencing data, unveiling associations between specific bacterial taxa and BV^16–19^. Beyond identifying microbial signatures, machine learning models have also been used to track the progression of BV and monitor treatment efficacy over time by identifying key microbial markers to predict BV recurrence with high accuracy^20^.

To harness these advancements and address challenges faced by women living with or at high risk for HIV, this study evaluates the accuracy of predictive models for diagnosing BV on a Tanzanian cohort of HIV positive women. We examine predictive accuracy using 16S rRNA gene sequence data and compare performance to two HIV-negative cohorts, highlighting the unique combination of bacterial communities important for accurate prediction. This research seeks to enhance diagnostic accuracy using advanced computational tools and highlight differences in microbial communities between groups.

## Methods

### Study population and design

Hummelen et al. studied and characterized the microbial communities of women living with HIV and investigated their relationship with BV diagnosis^17^. The study design and protocol were approved by the medical ethical review committee of Erasmus University Medical Centre, The Netherlands, and the medical research coordinating committee of the National Institute for Medical Research, Tanzania. The study was registered at clinical trials.gov NCT00536848. Participants were informed of the purpose of the study and provided their signed informed consent before participation. Between October 2007 and February 2008, HIV seropositive women attending the HIV care and treatment clinic at Sekou-Toure hospital were enrolled. This study resulted in a dataset consisting of 272 samples from 118 Tanzania women between 18 and 45 years old containing operational taxonomic unit (OTU) variables, total number of reads, Amsel’s criteria, and Nugent score. Of the 272 samples, 55% were positive for BV. The OTU variables were generated from sequencing the V6 region of the 16S rRNA gene obtained from vaginal swabs. Nugent score was indicated based on Gram stain test of vaginal smears, with 7-10 indicating a positive case, 4-6 intermediate, <3 BV negative. Prediction models were derived for diagnosing BV using the microbiome data from this HIV positive cohort of women in Tanzania.

Model predictive performance for diagnosing BV in HIV-positive cohort was compared to two HIV-negative cohorts (asymptomatic and symptomatic). The symptomatic cohort is produced from a study conducted by Srinivasan et al., which consists of 220 women with and without bacterial vaginosis (BV)^18^. The protocol was approved by the Institutional Review Board at the Fred Hutchinson Cancer Research Center. All study participants provided written informed consent. Participants were enrolled in the study between September 2006 and June 2010. BV was diagnosed based on Nugent score as described previously. Participants with BV were treated with intravaginal metronidazole gel each night for five days. Women were recruited from the Public Health, Seattle and King County Sexually Transmitted Diseases Clinic. This dataset is referred to as the symptomatic dataset.

The asymptomatic HIV-negative cohort was derived from a clinical study by Ravel et. al., which consists of 396 women with asymptomatic BV^12^. The study protocol was approved by the institutional review boards at Emory University School of Medicine, Grady Memorial Hospital, and the University of Maryland School of Medicine. The study was registered at clinicaltrials.govunder ID NCT00576797. Informed consent was obtained from participants. Participants were enrolled in the study between June 2008 and January 2009. The women were recruited from three clinical sites, two in Baltimore at the University of Maryland School of Medicine and one in Atlanta at Emory University. The women had the following characteristics: not pregnant, of reproductive age, ranging from 12 to 45 years, regularly menstruating, with a history of sexual activity, and had not taken any antibiotic or antimycotic compounds in the past 30 days. Exclusion criteria included using douches, vaginal medications or suppositories, feminine sprays, genital wipes or contraceptive spermicides, reported vaginal discharge in the past 48 hours, or menstruating or using contraceptives directly delivered to the vaginal mucosa at the time of the study. When referenced in this paper, this dataset is called the asymptomatic dataset. The clinical task for this study was to predict bacterial vaginosis (BV) from sequencing data for one HIV-positive and two HIV-negative cohorts. Predictive performance by cohort was compared to examine differences in model performance between HIV-positive and HIV-negative cohorts. We also examined differences in significant bacterial predictors. BV outcome was determined based on Nugent score, in which subjects with a Nugent score of seven or greater were labeled as BV positive and those with a score less than seven were labeled as BV negative.

### Predictors

For the Tanzanian, or HIV-positive cohort, we used 60 bacterial (OTU) variables as predictors of BV. For the HIV-negative cohorts, we used 247 and 155 OTU variables for the asymptomatic and symptomatic datasets, respectively. For all cohorts, the variables had no missing data. Predictor variables (i.e., OTU variables) were normalized by the total reads per sample, resulting in variable values between 0 and 1.

### Model development

Model development followed the Transparent Reporting of a multivariable prediction model for Individual Prognosis Or Diagnosis (TRIPOD+AI) guidelines^21^. We developed prediction models using machine learning and compared performances at predicting BV outcome between an HIV-positive cohort and two HIV-negative cohorts. We used random forest (RF), logistic regression (LR), support vector machine (SVM), and multi-layer perceptron (MLP) to predict BV outcome. We developed the models for each dataset using all bacterial variables available in each dataset. Predictive analyses were conducted in Python.

To minimize model overfitting, we performed a four-fold nested cross-validation, repeated 10 times. For each train-test fold split, we performed a cross-validated grid search within the train fold to find parameters for optimal model performance to train the model. Table 1 displays the hyperparameters tuned for each classifier. In the HIV cohort, some subjects have multiple samples; therefore, to avoid linkage between the train and test folds (i.e., data leakage), cross-validation was conducted with group stratification, which returned stratified folds with non-overlapping groups. The percentage of samples for each class (i.e., BV outcome) were preserved across each fold. In addition, each group (i.e., all samples belonging to a subject) appeared exactly once in the test set across all folds. For the HIV-negative cohorts, nested cross-validation was performed with stratification based on BV outcome and ethnicity. The percentage of samples for each class and ethnicity are preserved across each fold.

**Table 1.**
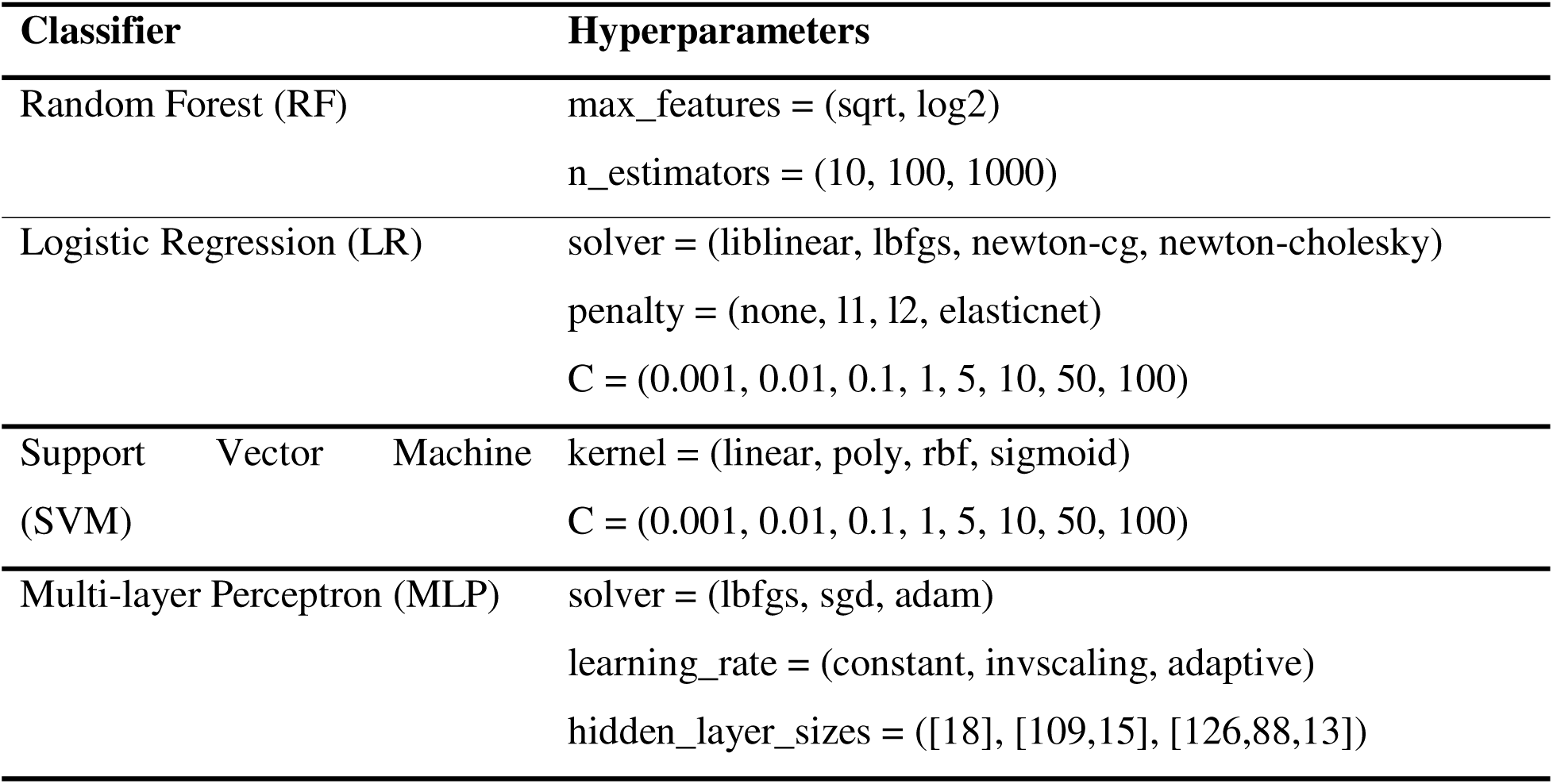
Hyperparameters for optimizing random forest (RF), logistic regression (LR), support vector machine (SVM), and multi-layer perceptron (MLP) models.

### Model evaluation

For the HIV-positive and HIV-negative cohorts, we evaluated model performance using the following metrics: <u>balanced accuracy (1), precision (2), recall (3), false positive rate (4), and</u> <u>false negative rate (5).</u> Below are the equations for computing each performance metric. Results were averaged across all folds from all runs to estimate the cross-validated model performance with 95% confidence intervals (CIs). We also evaluated FPR and FNR for Black women in the HIV-negative cohorts in comparison to the HIV-positive cohort, which consists of African women from Tanzania. In addition, permutation importance was performed to determine significant bacterial features.

Specifically for the HIV-positive cohort, we performed model calibration to investigate whether the predicted probabilities of BV outcome from the models closely matched the actual outcomes, specifically the percentage of BV-positive outcomes given the actual Nugent score. The sensitivity (6) and specificity (7) of each model was also examined. For specificity, we assessed the model’s accuracy for intermediate samples (i.e., samples with Nugent scores between four and six) and definite BV-negative samples (i.e., samples whose Nugent scores are three or less).

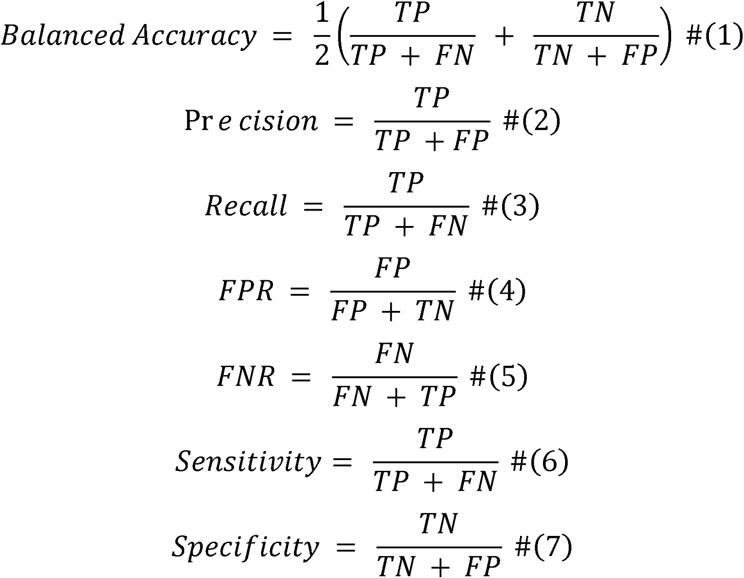

### Statistical analysis

To compare performance (i.e., balanced accuracy, precision, recall, FPR, and FNR) across the machine learning models, either a repeated measures, one-way ANOVA or Friedman’s statistical test was used, depending on whether the performance measures from the cross validation were normally distributed. Shapiro-Wilk test was used to determine normality. Post-hoc tests were conducted for an ANOVA or Friedman’s test that yield a significant result using multiple t-tests or a Wilcoxon-rank sum test, depending on data normality.

For comparing model performance between the BV cohorts, an independent measures, a one-way ANOVA or Kruskal-Wallis test was used given whether cross-validated measures were normally distributed.

All statistics were performed using “car”, “rstatix”, and “stats” packages in R (v4.3.1). Figures were created using Prism GraphPad (v10.4.0).

## Results

Predicting BV using sequencing data offers insights into the complexities of the vaginal microbiome, particularly for women of African descent, whose microbiomes exhibit greater variability even in healthy states. The choice of model architecture plays a crucial role in accurately capturing this variability and predicting BV outcomes. Therefore, we evaluated the effectiveness of four machine learning models, random forest (RF), logistic regression (LR), support vector machine (SVM), and multi-layer perceptron (MLP to understand how model architecture influences predictive performance within an HIV-positive cohort.

### Selection of optimal predictive model dependent on evaluation metric

Our findings reveal that no single model performed the best across all performance metrics (Figure 1, Table S1). Both Logistic Regression (LR) and Multi-Layer Perceptron (MLP) demonstrated the highest average AUROC scores (LR: 0.89, 95% CI: 0.87-0.90; MLP: 0.89, 95% CI: 0.88-0.91), indicating their effectiveness in distinguishing BV cases from negative cases. However, support vector machine (SVM) showed the lowest balanced accuracy (0.77, 95% CI: 0.75-0.79), suggesting it struggled to predict BV accurately within this cohort. Further analysis revealed that RF achieved the lowest false positive with an FPR of 0.19 (95% CI: 0.16-0.21). Interestingly, while SVM had the lowest balanced accuracy, it exhibited the highest average recall, correctly identifying 78% of true BV cases (95% CI: 0.75-0.82).

**Figure 1:**
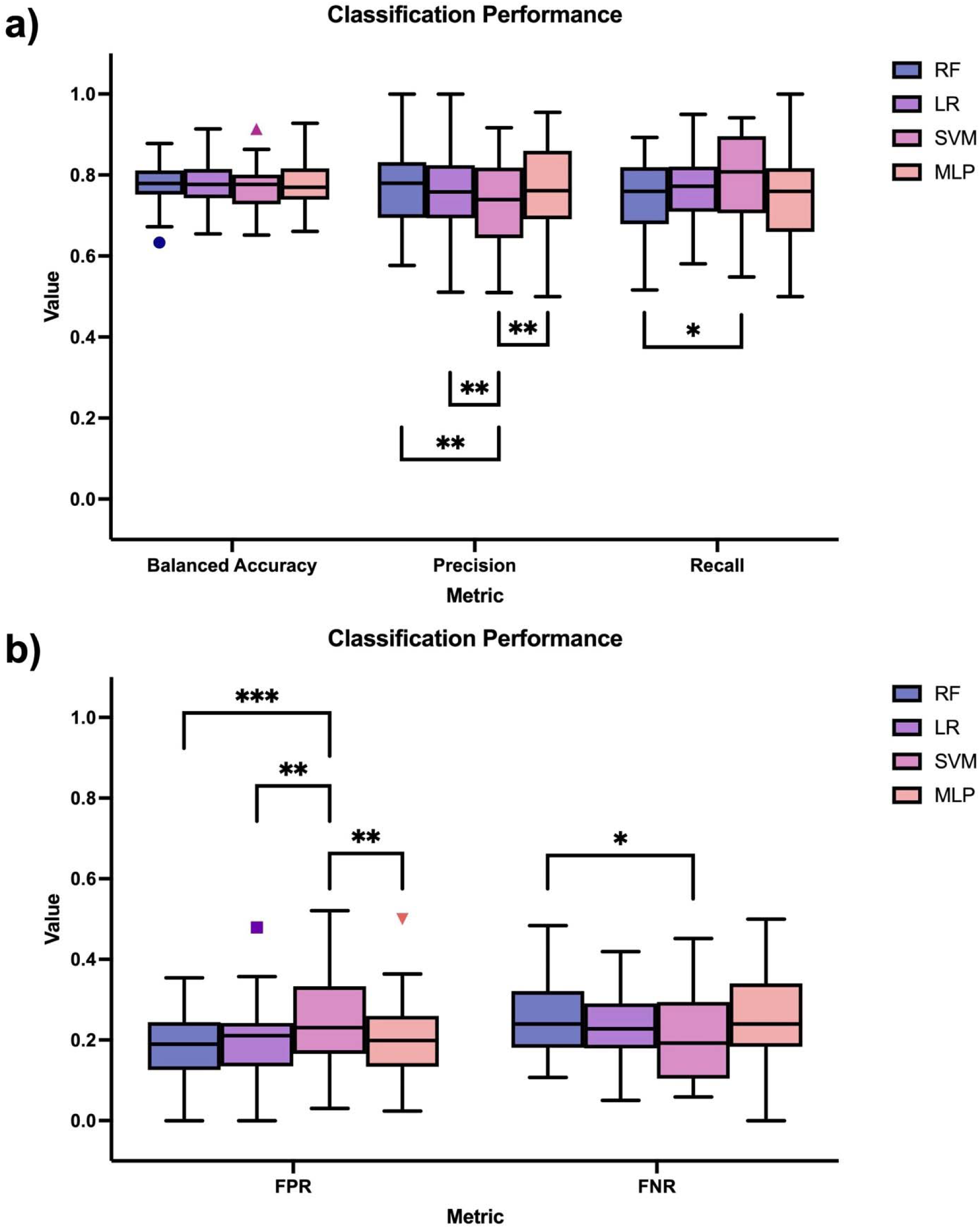
Performance score for predicting BV outcome using random forest (RF), logistic regression (LR), support vector machine (SVM), and multi-layer perceptron (MLP) classifiers. Boxplots showing the median, upper quartile, lower quartile, and outliers of a) balanced accuracy, precision, recall b) false positive rate (FPR), and false negative rate (FNR). Asterisk (*) denotes statistical significance in model performance between different BV outcomes.

### BV predictive performance lower for HIV+ cohort than HIV- cohorts

We compared the performance of machine learning models in this HIV-positive cohort against two HIV-negative cohorts to examine the influence of the vaginal microbiome communities on accurate prediction. Notably, the models were more accurate in predicting BV within the HIV-negative groups <u>(Tables S1-S3)</u>. False positive rates (FPR) were significantly higher in the HIV-positive cohort when compared to HIV-negative cohorts across all models (Figure 2a). False negative rates (FNR) were also higher in HIV-positive cohorts compared to HIV-negative cohorts, except when using Random Forest (Figure 2b).

**Figure 2.**
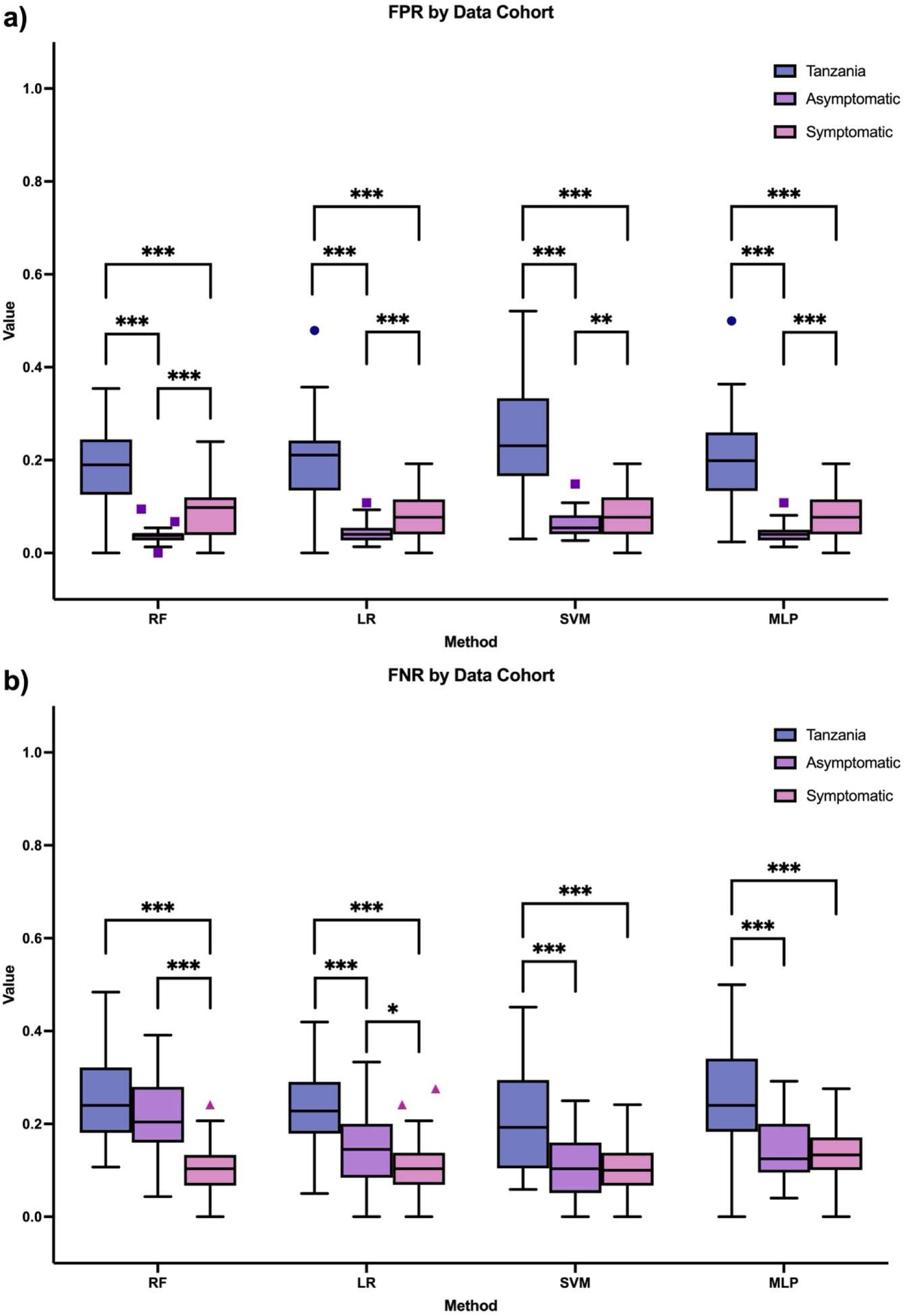
Performance score for predicting BV outcome in HIV and HIV-negative cohorts using random forest (RF), logistic regression (LR), support vector machine (SVM), and multi-layer perceptron (MLP) classifiers. Boxplots showing the median, upper quartile, lower quartile, and outliers of a) false positive rate (FPR) and b) false negative rate (FNR). Asterisk (*) denotes statistical significance in model performance between different BV outcomes.

Similar to the HIV-positive cohort, no single model performed the best across performance measures for asymptomatic and symptomatic cohorts. However, FPR tended to be lower for the asymptomatic dataset compared to the symptomatic dataset (Tables S2, S3). The opposite is true when examining FNR for these datasets.

### Influence of HIV status and microbial variation on model performance

HIV status has been found to impact vaginal microbiome composition^3,22,23^ To determine how this variation impacts model performance across the cohorts, we created two-dimensional (2-D) t-distributed stochastic Neighbor Embedding (t-SNE) projections of the sequence data for each cohort (Figure 3). Compared to the HIV-negative cohorts, the HIV-positive cohort had significant overlap across BV diagnosis clusters, particularly for subjects with intermediate Nugent scores (i.e., Nugent scores from 4 to 6). This overlap indicates a discrepancy in BV diagnosis where samples are not clearly classified, complicating accurate identification and treatment decisions. These correlate with the decrease in performance of various predictive models (RF, LR, SVM, MLP). Additionally, when observing bacterial composition by community state types (CST), most positive BV outcomes across all cohorts are associated with CST IV, in which no singular bacterial species is dominant but rather several different species make significant contributions to the vaginal microbiota. When examining BV negative outcomes, CST III is most prevalent with *L. iners* being the dominant bacterial species, CST I, which is *L. crispatus* dominant, was also associated with a negative diagnosis, more prevalently in the non-HIV cohorts. The HIV-negative cohorts also have some patients with *L. jensenii* dominant vaginal microbiome (CST V). Overall, the HIV-negative cohorts, tended to have *Lactobacilli*-dominant clusters (i.e., CST I and V) which are most optimal against BV, which were lacking in the Tanzania cohort.

**Figure 3.**
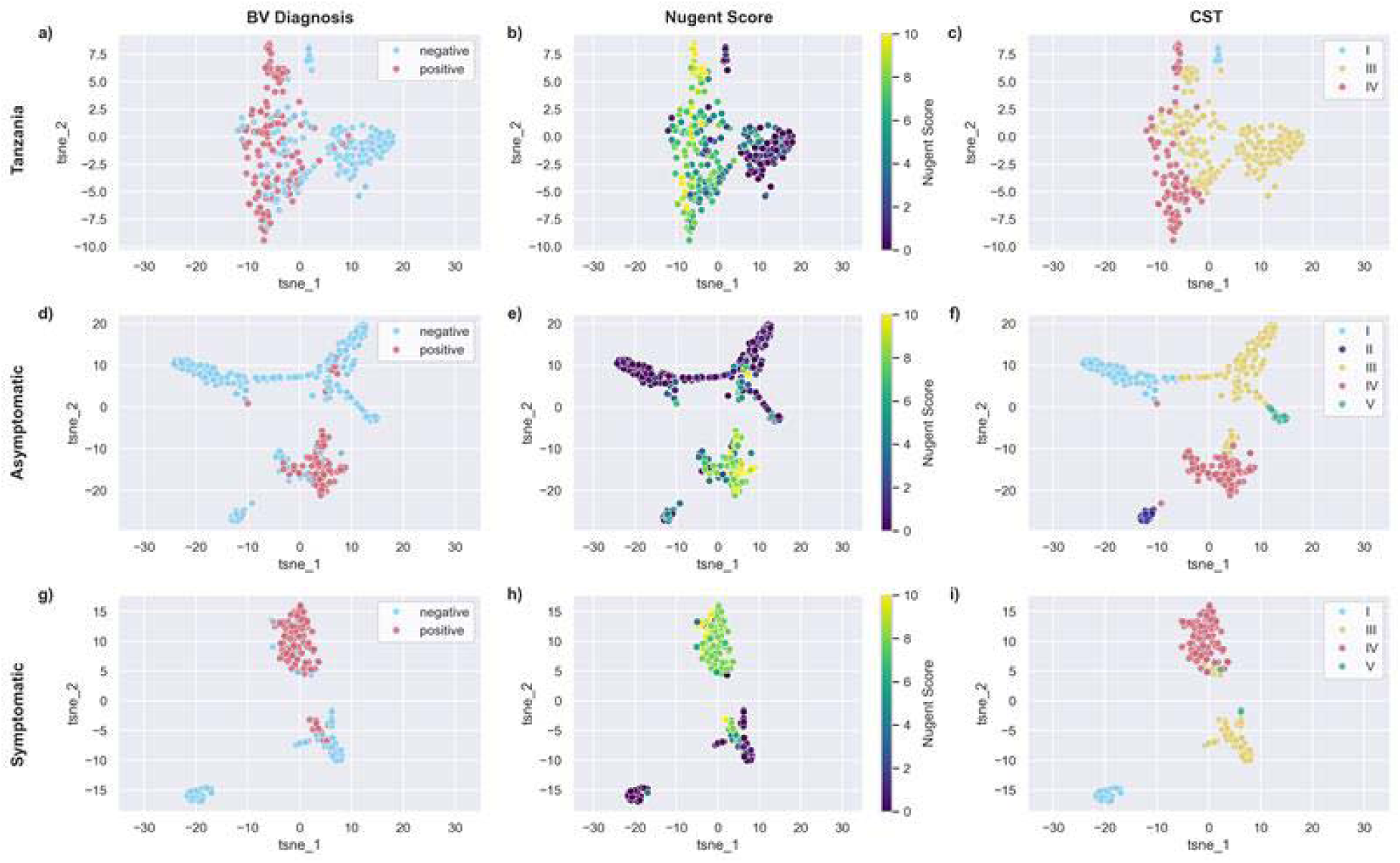
Two-dimensional t-SNE visualizations of bacterial taxa by BV diagnosis for three cohorts a) Tanzania (HIV+), d) Asymptomatic, and g) Symptomatic BV. Positive BV diagnoses are depicted in pink and negative BV diagnoses in blue. t-SNE visualizations of bacterial taxa by Nugent Score for b) Tanzania, e) Asymptomatic BV, and h) Symptomatic BV. Nugent score is depicted by the color palette t-SNE visualizations of bacterial taxa by community state type for c) Tanzania, f) Asymptomatic BV, and i) Symptomatic BV. CST I depicted in blue, II in purple, III in yellow, IV in pink, and V in green.

### Challenges in Predicting BV Across Intermediate and High Nugent Scores

Given that the HIV+ cohort displayed less clustering by BV status and intermediate Nugent scores often fall into a diagnostic gray area with a higher misclassification^24^, we investigated how accurately different machine learning architectures predict these ambiguous cases within the HIV-positive cohort. By examining predictive outcomes based on Nugent scores (Figure 4, Table 2), we found that although samples with intermediate scores (4 to 6) were labeled as BV negative, a significant portion was incorrectly predicted as BV positive across all models. Random Forest (RF) performed best at accurately classifying intermediate samples as BV negative, followed by the Multi-Layer Perceptron (MLP). While models were generally more adept at predicting samples with low Nugent scores (0-3) as BV negative, they consistently identified samples with high Nugent scores (7-10) as BV positive (Table 2). Notably, SVM was the most effective at predicting BV positive for high Nugent scores, whereas Logistic Regression (LR) and MLP excelled at predicting BV negative for samples with low Nugent scores.

**Figure 4.**
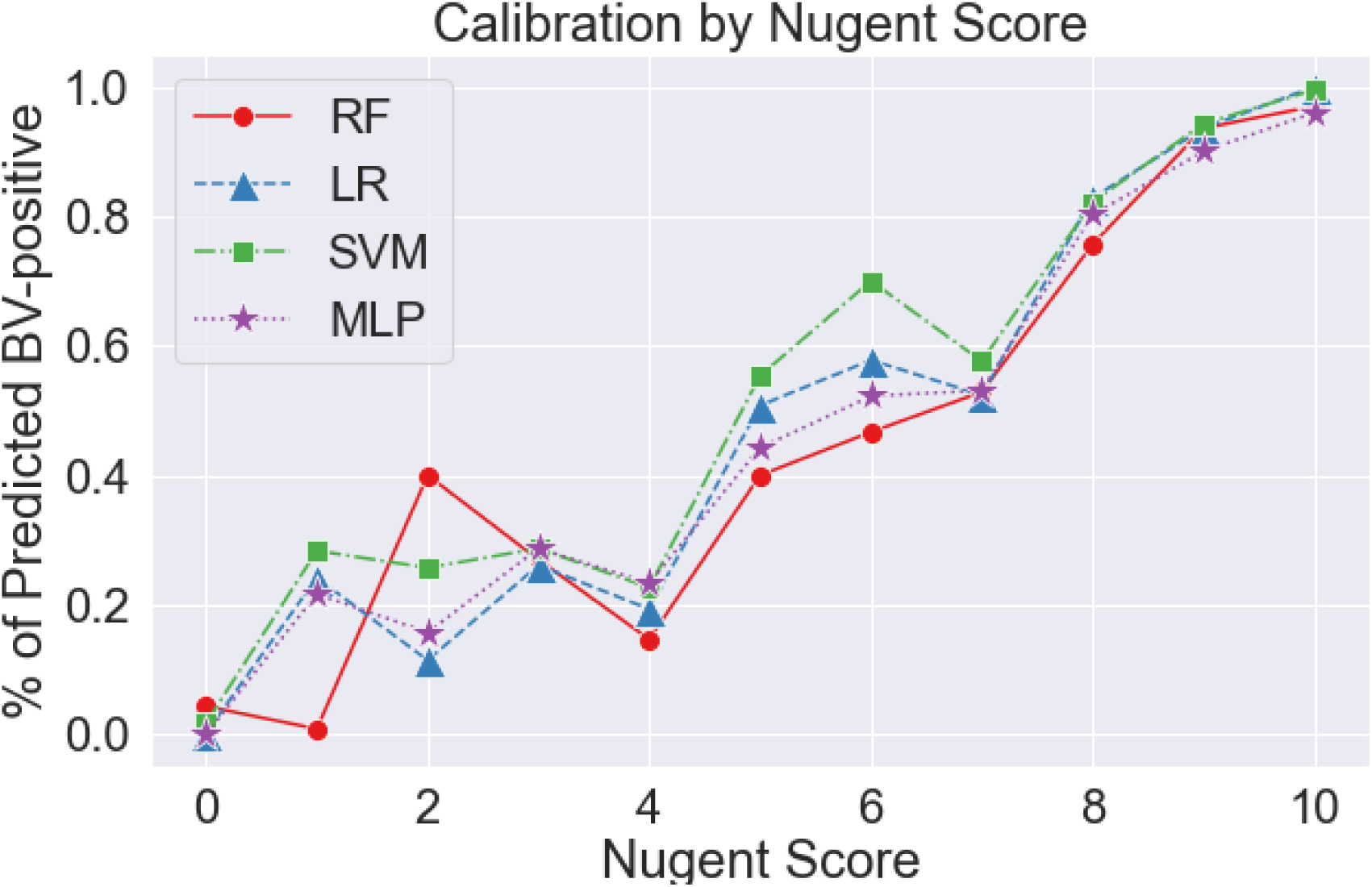
Fraction of BV-positive outcome at a given Nugent score for logistic regression (LR), random forest (RF), support vector machine (SVM), and multi-layer perceptron (MLP) models.

**Table 2.** Percentage correctly labelled for each BV outcome – positive (7-10), intermediate (4-6), negative (0-3) - based on Nugent score from random forest (RF), logistic regression (LR), support vector machine (SVM), and multi-layer perceptron (MLP) classifiers. Since performing binary classification, BV-intermediates were labelled as BV-negative (i.e., percentage of BV-intermediate samples correctly labelled as BV-negative). □

|  | RF | LR | SVM | MLP |
| --- | --- | --- | --- | --- |
| BV-positive | 0.74 | 0.77 | 0.78 | 0.75 |
| BV-intermediate | **0.67** | **0.59** | **0.52** | **0.61** |
| BV-negative | 0.88 | 0.90 | 0.87 | 0.90 |

### Key predictors of BV across HIV-positive and HIV-negative Cohorts

To determine how variations in bacterial composition influence model performance between th HIV-positive and HIV-negative cohorts, we compared the most significant predictors of BV across each cohort. The top eight significant features from each cohort are displayed below (Table 3). Some overlap was observed between the HIV-positive and HIV-negative cohorts, specifically with *Prevotella*, *Gardnerella vaginalis*, and *Dialister*. However, the top most significant predictor of BV for each cohort differ with *L. iners* being the most important bacterial taxa among the Tanzania cohort, *Parvimonas micra* for the symptomatic BV cohort, and *Gardnerella* for the asymptomatic BV cohort.

**Table 3.** Important predictor variables (i.e., bacterial, or operational taxonomic unit, features) for predicting bacterial vaginosis in HIV (i.e., Tanzania) and HIV-negative cohorts (i.e., symptomatic and asymptomatic) cohorts using random forest (RF) model.

| Tanzania□ | Symptomatic | Asymptomatic |
| --- | --- | --- |
| L. iners□ | Parvimonas micra□ | Gardnerella□ |
| Prevotella timonensis<sup>b</sup>□ | Gardnerella vaginalis<sup>b</sup>□ | Dialister□ |
| Gardnerella vaginalis<sup>b</sup>□ | Atopobium vaginae<sup>b</sup>□ | Prevotella□□ |
| Dialister micraerophilus□ | Prevotella timonensis<sup>b</sup>□ | L. crispatus<sup>b</sup>□ |
| Veillonella montpellierensis□ | Megasphaera sp. type 1<sup>□</sup>□ | Peptostreptococcus□ |
| Prevotella amniotica<sup>□</sup>□ | L. crispatus<sup>b</sup>□ | Actinomyces□ |
| Atopobium vaginae<sup>b</sup>□ | Lactobacillus vaginalis<sup>□</sup>□ | Peptoniphilus□ |
| Campylobacter□ | Dialister sp. type 2□ | Veillonella<sup>□</sup>□ |

### Model Performance for Black Women Across HIV-Positive and HIV-Negative Cohorts

Given women of African ancestry tend to have a more diverse vaginal microbiota, we compared model performance of Black women across all cohorts. Similarly, we found that models performed worst for the HIV-positive cohort compared to Black women from the symptomatic and asymptomatic (HIV-negative) cohorts (Figure 5b; Tables S4-S5), except when examining false positive rate (FPR). The false positive rates for predicting BV in the HIV-positive cohort and symptomatic (HIV-negative) cohort were similar for most models (Figure 5a).□□

**Figure 5.**
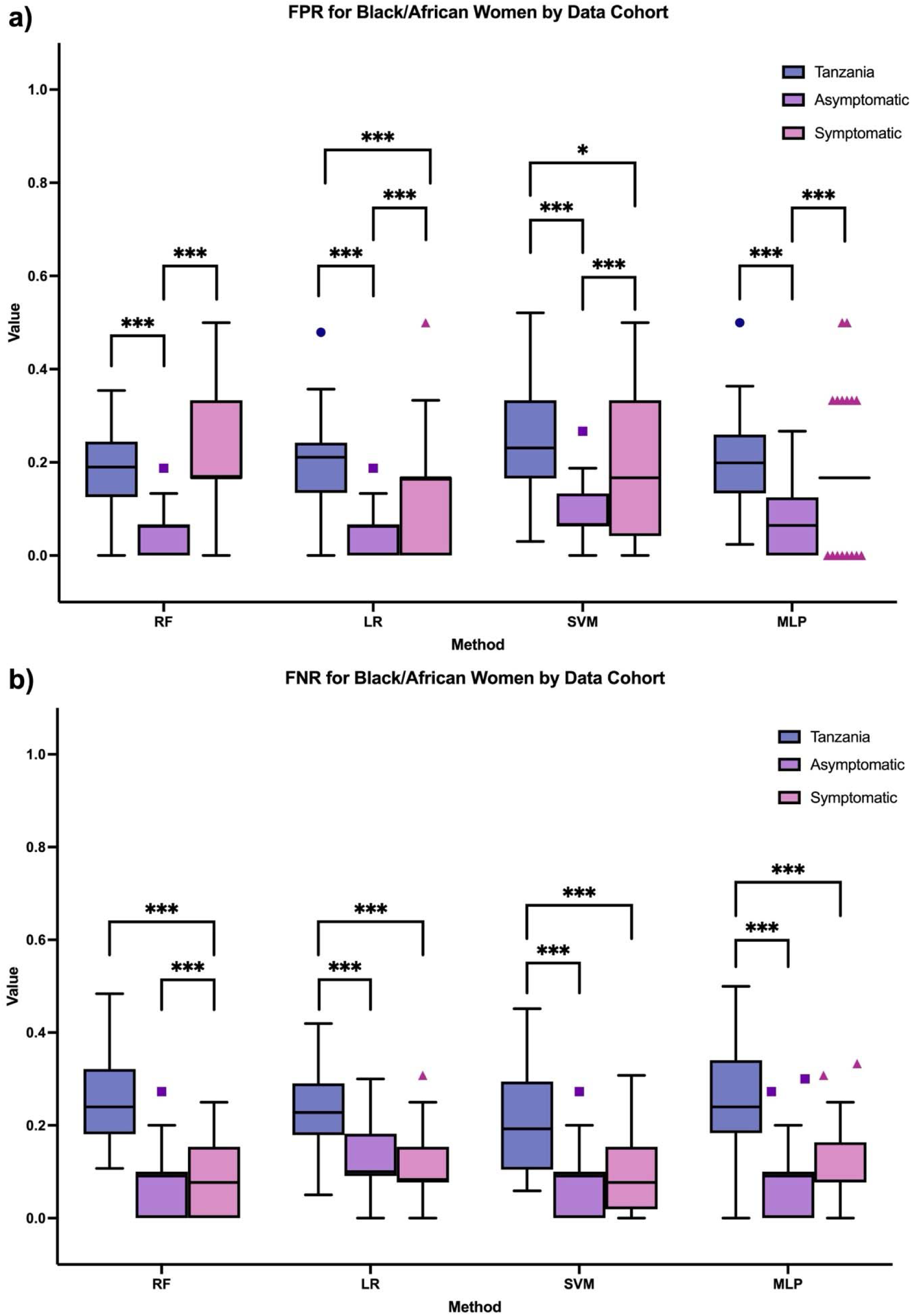
Performance score for predicting BV outcome for Black women in HIV and non-HIV cohorts using random forest (RF), logistic regression (LR), support vector machine (SVM), and multi-layer perceptron (MLP) classifiers. Boxplots showing the median, upper quartile, lower quartile, and outliers of a) false positive rate (FPR) and b) false negative rate (FNR). Asterisk (*) denotes statistical significance in model performance between different BV outcomes.

## Discussion

HIV and BV disproportionately impact women of African descent. Women of color tend to have a more diverse vaginal microbiome, even in healthy states, which can lead to discrepancies in BV diagnosis. Additionally, prior work has found that HIV status influences vaginal microbiome composition. We evaluate the difference in model performance of four machine learning classifiers for diagnosing BV between a HIV-positive cohort of Tanzanian women and two HIV-negative cohorts of women from the United States. Overall, models tended to perform similarly in predicting BV within the HIV-positive cohort. However, model performance for the Tanzanian cohort was significantly lower compared to the HIV-negative cohorts (Figure 2). The use of t-SNE projection of OTU variables mapped by positive and negative BV diagnosis allowed for visualization of taxonomic clusters. Data from Tanzanian dataset was very distinctly distributed compared to the other datasets. Additionally, when comparing model performance across cohorts for African/Black women, models performed significantly better for Black women (i.e., within United States) from the HIV-negative cohorts than the Tanzanian women from the HIV-positive cohort (Figure 5). This comparative analysis of the vaginal microbiome has exposed potential inequities in BV predictive outcomes for women living with HIV.

While models tended to perform similarly in predicting BV the Tanzanian cohort, model performance for this HIV-positive cohort was significantly lower than for the HIV-negative cohorts (Figure 4). Prior works have demonstrated the feasibility of using machine learning models and vaginal 16S rRNA sequencing data to indicate BV diagnosis using the two HIV-negative cohorts (i.e., asymptomatic and symptomatic BV datasets)^16,17,25,26^. Similar to prior studies, both symptomatic^18^ and asymptomatic^12^ datasets had similar predictive accuracy (i.e., balanced accuracy, AUROC, and FNR), despite amplifying different hypervariable regions, V3-V4 and V1-V2, respectively. Additionally, each study used different methods for classifying reads into taxonomic groups (i.e., RDP Naive Bayesian classifier in asymptomatic^12^, placer in symptomatic^27^). The sequence data for the HIV-positive cohort in this study was obtained from the V6 region and used UCLUST for classification ^28^. As previous differences in sequence region and taxonomic classification method did not significantly affect results for predicting BV in HIV-negative datasets, we hypothesize that differences in predictive performance between the cohorts may be due to differences in HIV status. However, the use of only one hypervariable region in the Tanzanian dataset may also pose challenges.

Further analysis in our study revealed the potential influence of intermediate Nugent scores in misdiagnoses of BV among the HIV-positive cohort (Table 2, Figures 3 and 4), specifically misclassifying intermediate Nugent samples as BV-positive. While traditionally considered BV-negative or excluded in studies, intermediate Nugent scores compromise the cervicovaginal mucus barrier to HIV similarly to high Nugent scores (7-10)^29^. This indicates that intermediate scores increase susceptibility to HIV infection due to their association with *L. iners-dominant* bacterial community. *L. iners* is found to be the least protective *lactobacilli* species for BV and sexually transmitted infections due to its characteristic of coexisting with both healthy and disruptive bacteria^30^, making it more likely to shift in and out of healthy states. Our findings suggest that including intermediate scores in BV diagnosis could enhance sensitivity and reveal the underestimated risk of HIV.

Upon examining significant predictors of BV for each cohort, we discovered few shared variables between the cohorts, further highlighting the differences between the HIV and HIV-negative cohorts. *L. iners* was found to be the most significant variable in indicating BV for the Tanzanian cohort. Based on our t-SNE projections (Figure 3c, 3f, and 3i), the presence of *L. iners* in most samples may have contributed to significant discrepancies in BV diagnosis. In contrast, the top predictors of BV for the HIV-negative cohorts were *Parvimonas micra* and *Gardnerella*, which are both anerobic bacteria. The presence of anaerobic bacteria, or lack thereof, is often an indicator of BV status, and may be a contributing factor to relatively higher predictive performance. Overall, the HIV cohort shared the most significant predictors with the symptomatic cohort (i.e., *Prevotella timonensis*, *Gardnerella vaginalis*, and *Atopobium vaginae*), although resulting in different predictive performance.

While our work revealed significant findings, there were also some limitations in conducting this work. As previously mentioned, sequence data for each cohort was extracted from different hypervariable regions, which may have contributed to differences in model performance and require further investigation. Additionally, our cohorts only represented women in the United States and Tanzania, which prevents generalization of our findings to cohorts containing samples of women of other ethnic backgrounds.

## Conclusions

The vaginal microbiome has mostly been studied in the context of HIV prevention^4,31–33^, and the impact of HIV status on microbial composition is understudied. This study thoroughly investigates the potential of machine learning models to predict BV in an HIV-positive cohort using 16S rRNA gene sequence data and compares these predictions to those for HIV-negative cohorts. Comparing model performance between the HIV-positive and HIV-negative cohorts shows that while the machine learning models perform well overall, their predictive accuracy is subtly affected by the cohort’s HIV status. We also identify bacterial components important to consider for vaginal health in HIV positive women. The identification of BV predictors from bacterial OTU taxa will inform future development of targeted therapeutics for BV for women living with HIV.

## Authors’ contributions

DPO, WG, and IKP designed the study. DPO designed machine learning analysis and experimental setup. DPO and CC prepared figures. CC conducted the statistical analysis. IKP supervised the project. All authors contributed to the initial draft of the manuscript. All authors contributed critical revisions to the manuscript and approved the final manuscript.

## Acknowledgements

We would like to acknowledge all women who provided samples for the data used in this manuscript.

## Data Availability Statement

The bacterial 16S rRNA gene sequences for the asymptomatic and symptomatic datasets analyzed during the current study are available in the National Center for Biotechnology Information Short Read Archive (SRA022855; <u>SRA051298</u>). Data on race and ethnicity for the HIV-negative datasets can found at the following links: [https://www.pnas.org/doi/full/10.1073/pnas.1002611107], and [https://journals.plos.org/plosone/article?id=10.1371/journal.pone.0037818#s5]. For the Tanzanian cohort, the bacterial 16S rRN gene sequences are available as supporting information within the publication (https://journals.plos.org/plosone/article?id=10.1371/journal.pone.0012078).

The code used to run experiments and generate results is available on GitHub at [https://github.com/The-Parker-Lab/Tanzania-HIV-BV-ML], including a README file explaining the details of the repository and how to run the code.

## Supporting Information

Supporting Information file 1: Additional tables of results from BV predictive analysis

Provides numerical values of model performance for predicting BV across the HIV and HIV-negative cohorts, both overall and for Black or African women within the HIV-negative cohorts. Additional tables providing top predictor variables of BV for the HIV-negative cohorts.

## References

1. The Lancet Hiv. Women lead the way in the HIV response. The Lancet HIV 11, e131 (2024).

2. The path that ends AIDS: UNAIDS Global AIDS Update 2023. Geneva: Joint United Nations Programme on HIV/AIDS; 2023. Licence: CC BY-NC-SA 3.0 IGO.

3. Atashili, J., Poole, C., Ndumbe, P. M., Adimora, A. A. & Smith, J. S. Bacterial vaginosis and HIV acquisition: a meta-analysis of published studies. AIDS 22, 1493–1501 (2008).

4. Chen, X., Lu, Y., Chen, T. & Li, R. The Female Vaginal Microbiome in Health and Bacterial Vaginosis. Front. Cell. Infect. Microbiol. 11, 631972 (2021).

5. Leitich, H. et al. Bacterial vaginosis as a risk factor for preterm delivery: A meta-analysis. American Journal of Obstetrics and Gynecology 189, 139–147 (2003).

6. Guidelines for the Management of Symptomatic Sexually Transmitted Infections. (World Health Organization, Geneva, 2021).

7. V Majigo, M., Kashindye, P., Mtulo, Z. & Joachim, A. Bacterial vaginosis, the leading cause of genital discharge among women presenting with vaginal infection in Dar es Salaam, Tanzania. Afr H. Sci. 21, 531–537 (2021).

8. Anahtar, M. N. et al. Cervicovaginal Bacteria Are a Major Modulator of Host Inflammatory Responses in the Female Genital Tract. Immunity 42, 965–976 (2015).

9. Amsel, R. et al. Nonspecific vaginitis. The American Journal of Medicine 74, 14–22 (1983).

10. Nugent, R. P., Krohn, M. A. & Hillier, S. L. Reliability of diagnosing bacterial vaginosis is improved by a standardized method of gram stain interpretation. J Clin Microbiol 29, 297–301 (1991).

11. Fettweis, J. M. et al. The vaginal microbiome and preterm birth. Nat Med 25, 1012–1021 (2019).

12. Ravel, J. et al. Vaginal microbiome of reproductive-age women. Proc. Natl. Acad. Sci. U.S.A. 108, 4680–4687 (2011).

13. Onywera, H., Williamson, A.-L., Mbulawa, Z. Z. A., Coetzee, D. & Meiring, T. L. Factors associated with the composition and diversity of the cervical microbiota of reproductive-age Black South African women: a retrospective cross-sectional study. PeerJ 7, e7488 (2019).

14. Peters, D. H., et al. Poverty and Access to Health Care in Developing Countries. Annals of the New York Academy of Sciences 1136, 161–171 (2008).

15. Peprah, P. et al. Lessening barriers to healthcare in rural Ghana: providers and users’ perspectives on the role of mHealth technology. A qualitative exploration. BMC Med Inform Decis Mak 20, 27 (2020).

16. Celeste, C. et al. Ethnic disparity in diagnosing asymptomatic bacterial vaginosis using machine learning. *npj Digit*. Med. 6, 211 (2023).

17. Beck, D. & Foster, J. A. Machine Learning Techniques Accurately Classify Microbial Communities by Bacterial Vaginosis Characteristics. PLoS ONE 9, e87830 (2014).

18. Beck, D. & Foster, J. A. Machine learning classifiers provide insight into the relationship between microbial communities and bacterial vaginosis. BioData Mining 8, 23 (2015).

19. Ma, Z. (Sam). A new hypothesis on BV etiology: dichotomous and crisscrossing categorization of complex versus simple on healthy versus BV vaginal microbiomes. mSystems 8, e00049–23 (2023).

20. Challa, A. et al. Multi□omics analysis identifies potential microbial and metabolite diagnostic biomarkers of bacterial vaginosis. Acad Dermatol Venereol 38, 1152–1165 (2024).

21. Collins, G. S. et al. TRIPOD+AI statement: updated guidance for reporting clinical prediction models that use regression or machine learning methods. BMJ e078378 (2024) doi:10.1136/bmj-2023-078378.

22. Nunn, K. L. et al. Enhanced Trapping of HIV-1 by Human Cervicovaginal Mucus Is Associated with Lactobacillus crispatus-Dominant Microbiota. mBio 6, e01084–15 (2015).

23. Mirmonsef, P. et al. A Comparison of Lower Genital Tract Glycogen and Lactic Acid Levels in Women and Macaques: Implications for HIV and SIV Susceptibility. AIDS Research and Human Retroviruses 28, 76–81 (2012).

24. Coleman, J. S. & Gaydos, C. A. Molecular Diagnosis of Bacterial Vaginosis: an Update. J Clin Microbiol 56, e00342–18 (2018).

25. Baker, Y. S., Agrawal, R., Foster, J. A., Beck, D. & Dozier, G. APPLYING MACHINE LEARNING TECHNIQUES IN DETECTING BACTERIAL VAGINOSIS. Proc Int Conf Mach Learn Cybern 2014, 241–246 (2014).

26. Pérez-Gómez, J. F., Canul-Reich, J., Hernández-Torruco, J. & Hernández-Ocaña, B. Predictor Selection for Bacterial Vaginosis Diagnosis Using Decision Tree and Relief Algorithms. Applied Sciences 10, 3291 (2020).

27. Srinivasan, S. et al. Bacterial Communities in Women with Bacterial Vaginosis: High Resolution Phylogenetic Analyses Reveal Relationships of Microbiota to Clinical Criteria. PLoS ONE 7, e37818 (2012).

28. Hummelen, R. et al. Deep Sequencing of the Vaginal Microbiota of Women with HIV. PLoS ONE 5, e12078 (2010).

29. Amabebe, E. & Anumba, D. O. C. The Vaginal Microenvironment: The Physiologic Role of Lactobacilli. Front. Med. 5, 181 (2018).

30. Petrova, M. I., Reid, G., Vaneechoutte, M. & Lebeer, S. Lactobacillus iners□: Friend or Foe? Trends in Microbiology 25, 182–191 (2017).

31. McClelland, R. S. et al. Evaluation of the association between the concentrations of key vaginal bacteria and the increased risk of HIV acquisition in African women from five cohorts: a nested case-control study. Lancet Infect Dis 18, 554–564 (2018).

32. Alisoltani, A. et al. Microbial function and genital inflammation in young South African women at high risk of HIV infection. Microbiome 8, 165 (2020).

33. Parker, I. K. et al. A multiplex assay for detection of SHIV plasma and mucosal IgG and IgA. Journal of Immunological Methods 450, 34–40 (2017).

