## Supplemental Figures and Tables for "Predictive modeling for bacterial vaginosis in a Tanzanian cohort of women living with HIV"

**Supporting Information.**

**Table S1. Classification performance for predicting bacterial vaginosis for HIV cohort using random forest (RF), logistic regression (LR), support vector machine (SVM), and multi-layer perceptron (MLP) models. Balanced accuracy (BACC), precision, recall, false positive rate (FPR), and false negative rate (FNR) provided with 95% confidence interval.**

| METRIC | RF | LR | SVM | MLP |
| --- | --- | --- | --- | --- |
| AUROC | 0.87 [0.86, 0.89] | **0.89 [0.87, 0.90]** | 0.88 [0.86, 0.89] | **0.89 [0.88, 0.91]** |
| BACC | **0.78 [0.76, 0.79]** | **0.78 [0.77, 0.80]** | 0.77 [0.75, 0.79] | **0.78 [0.76, 0.8]** |
| Precision | **0.77 [0.73, 0.80]** | 0.76 [0.72, 0.79] | 0.73 [0.69, 0.76] | 0.76 [0.73, 0.80] |
| Recall | 0.74 [0.71, 0.77] | 0.77 [0.74, 0.79] | **0.78 [0.75, 0.82]** | 0.75 [0.72, 0.79] |
| FPR | **0.19 [0.16, 0.21]** | 0.20 [0.18, 0.23] | 0.24 [0.21, 0.28] | 0.20 [0.16, 0.23] |
| FNR | 0.26 [0.23, 0.29] | 0.23 [0.21, 0.26] | **0.22 [0.18, 0.25]** | 0.25 [0.21, 0.28] |

**Table S2. Classification performance for predicting bacterial vaginosis for HIV-negative, symptomatic BV cohort using random forest (RF), logistic regression (LR), support vector machine (SVM), and multi-layer perceptron (MLP) models. Balanced accuracy (BACC), precision, recall, false positive rate (FPR), and false negative rate (FNR) provided with 95% confidence interval.**

| METRIC | RF | LR | SVM | MLP |
| --- | --- | --- | --- | --- |
| AUROC | **0.97 [0.96, 0.97]** | 0.95 [0.95, 0.96] | 0.95 [0.94, 0.96] | 0.95 [0.94, 0.96] |
| BACC | **0.90 [0.89, 0.91]** | **0.90 [0.89, 0.91]** | **0.90 [0.89, 0.91]** | 0.89 [0.88, 0.90] |
| Precision | 0.92 [0.91, 0.93] | 0.92 [0.91, 0.94] | 0.92 [0.90, 0.93] | 0.92 [0.91, 0.93] |
| Recall | **0.90 [0.89, 0.92]** | 0.88 [0.87, 0.90] | **0.90 [0.88, 0.92]** | 0.87 [0.85, 0.89] |
| FPR | 0.09 [0.08, 0.11] | 0.09 [0.07, 0.10] | 0.09 [0.08, 0.11] | 0.09 [0.07, 0.10] |
| FNR | **0.10 [0.08, 0.11]** | 0.12 [0.10, 0.13] | **0.10 [0.08, 0.12]** | 0.13 [0.11, 0.15] |

**Table S3. Classification performance for predicting bacterial vaginosis for HIV-negative, asymptomatic BV cohort using random forest (RF), logistic regression (LR), support vector machine (SVM), and multi-layer perceptron (MLP) models. Balanced accuracy (BACC), precision, recall, false positive rate (FPR), and false negative rate (FNR) provided with 95% confidence interval.**

| METRIC | RF | LR | SVM | MLP |
| --- | --- | --- | --- | --- |
| AUROC | 0.96 [0.96, 0.97] | **0.97 [0.96, 0.97]** | 0.96 [0.95, 0.96] | 0.96 [0.95, 0.96] |
| BACC | 0.87 [0.86, 0.89] | 0.90 [0.89, 0.92] | **0.91 [0.9, 0.92]** | **0.91 [0.9, 0.92]** |
| Precision | **0.88 [0.86, 0.89]** | 0.87 [0.86, 0.89] | 0.83 [0.81, 0.85] | **0.88 [0.87, 0.9]** |
| Recall | 0.78 [0.76, 0.81] | 0.85 [0.83, 0.87] | **0.89 [0.87, 0.91]** | 0.85 [0.83, 0.87] |
| FPR | **0.04 [0.03, 0.04]** | **0.04 [0.04, 0.05]** | 0.06 [0.05, 0.07] | **0.04 [0.03, 0.04]** |
| FNR | 0.22 [0.19, 0.24] | 0.15 [0.13, 0.17] | **0.11 [0.09, 0.13]** | 0.15 [0.13, 0.17] |

**Table S4. BV predictive performance for Black women from the HIV-negative, symptomatic BV cohort using random forest (RF), logistic regression (LR), support vector machine (SVM), and multi-layer perceptron (MLP) models. Balanced accuracy (BACC), precision, recall, false positive rate (FPR), and false negative rate (FNR) provided with 95% confidence interval.**

| METRIC | RF | LR | SVM | MLP |
| --- | --- | --- | --- | --- |
| AUROC | **0.95 [0.93, 0.96]** | 0.92 [0.90, 0.95] | 0.92 [0.89, 0.94] | 0.93 [0.91, 0.95] |
| BACC | 0.85 [0.83, 0.88] | **0.87 [0.85, 0.89]** | 0.86 [0.84, 0.88] | 0.85 [0.83, 0.88] |
| Precision | 0.91 [0.89, 0.93] | **0.93 [0.91, 0.95]** | 0.92 [0.90, 0.94] | 0.92 [0.90, 0.93] |
| Recall | **0.92 [0.89, 0.94]** | 0.88 [0.86, 0.91] | 0.91 [0.88, 0.93] | 0.88 [0.85, 0.90] |
| FPR | 0.21 [0.16, 0.25] | **0.14 [0.10, 0.18]** | 0.18 [0.14, 0.23] | 0.18 [0.14, 0.21] |
| FNR | **0.08 [0.06, 0.11]** | 0.12 [0.09, 0.14] | 0.09 [0.07, 0.12] | 0.12 [0.10, 0.15] |

**Table S5. BV predictive performance for Black women from the HIV-negative, asymptomatic BV cohort using random forest (RF), logistic regression (LR), support vector machine (SVM), and multi-layer perceptron (MLP) models. Balanced accuracy (BACC), precision, recall, false positive rate (FPR), and false negative rate (FNR) provided with 95% confidence interval.**

| METRIC | RF | LR | SVM | MLP |
| --- | --- | --- | --- | --- |
| AUROC | **0.98 [0.97, 0.99]** | 0.97 [0.96, 0.98] | 0.96 [0.95, 0.97] | 0.97 [0.97, 0.98] |
| BACC | **0.93 [0.92, 0.94]** | 0.92 [0.9, 0.93] | 0.92 [0.91, 0.94] | 0.92 [0.9, 0.94] |
| Precision | **0.93 [0.91, 0.95]** | 0.92 [0.9, 0.95] | 0.89 [0.87, 0.91] | 0.91 [0.88, 0.93] |
| Recall | 0.92 [0.89, 0.94] | 0.89 [0.86, 0.91] | **0.93 [0.9, 0.95]** | 0.91 [0.88, 0.93] |
| FPR | **0.05 [0.04, 0.07]** | **0.05 [0.04, 0.07]** | 0.08 [0.06, 0.1] | 0.07 [0.05, 0.09] |
| FNR | 0.08 [0.06, 0.11] | 0.11 [0.09, 0.14] | **0.07 [0.05, 0.1]** | 0.09 [0.07, 0.12] |
